# Associations between neuromelanin depletion and cortical rhythmic activity in Parkinson’s disease

**DOI:** 10.1101/2024.02.16.24302958

**Authors:** Alex I. Wiesman, Victoria Madge, Edward A. Fon, Alain Dagher, D Louis Collins, Sylvain Baillet, PREVENT-AD Research Group, Quebec Parkinson Network

## Abstract

**Background and Objectives:** Parkinson’s disease (PD) is marked by the death of neuromelanin-rich dopaminergic and noradrenergic cells in the substantia nigra (SN) and the locus coeruleus (LC), respectively, resulting in motor and cognitive impairments. While SN dopamine dysfunction has clear neurophysiological effects, the impact of reduced LC norepinephrine signaling on brain activity in PD remains to be established.

**Methods:** We used neuromelanin-sensitive T1-weighted MRI (N_PD_ = 58; N_HC_ = 27) and task-free magnetoencephalography (N_PD_ = 58; N_HC_ = 65) to identify neuropathophysiological factors related to the degeneration of the LC and SN in patients with PD.

**Results:** We found pathological increases in rhythmic alpha (8 – 12 Hz) activity in patients with decreased LC neuromelanin, with a stronger association in patients with worse attentional impairments. This negative alpha–LC neuromelanin relationship is also stronger in fronto-motor cortices, which are regions with high densities of norepinephrine transporters in the healthy brain, and where alpha activity is negatively related to attention scores. These observations support a noradrenergic association between LC integrity and alpha band activity. Our data also show that rhythmic beta (15 – 29 Hz) activity in the left somato-motor cortex decreases with lower levels of SN neuromelanin; the same regions where beta activity reflects axial motor symptoms.

**Discussion:** Together, our findings clarify the association of well-documented alterations of rhythmic neurophysiology in PD with cortical and subcortical neurochemical systems. Specifically, attention-related alpha activity reflects dysfunction of the noradrenergic system, and beta activity with relevance to motor impairments reflects dopaminergic dysfunction.

## Introduction

One of the defining histopathological features of Parkinson’s disease (PD) is the loss of neuromelanin-rich cells in brainstem nuclei. Degeneration of such dopaminergic neurons in the substantia nigra (SN) leads to dysfunctional signaling along the subcortical nigrostriatal pathway and contributes to motor impairments^1^. Loss of neuromelanin-rich neurons in the locus coeruleus (LC) can precede SN deterioration^2^ and disrupts the production of norepinephrine in the central nervous system. The resulting changes in noradrenergic signalling projecting to the cortex are thought to induce cognitive symptoms^3^, primarily impairment of attention functions^4, 5^.

Novel neuromelanin-sensitive MRI measurements of SN and LC integrity hold promise for the non-invasive monitoring of the degeneration of dopaminergic and noradrenergic systems in PD^6, 7^ in relation to the respective hallmark motor and cognitive features of the disease^3, 7^. A knowledge gap remains between these neurochemical observations and the well-documented alterations of rhythmic and arrhythmic neurophysiological activity in PD^8–10^, and whether any such associations relate to the cognitive and motor features of PD.

In particular, cortical alpha rhythms (8 – 12 Hz) are altered in PD, and, akin to the deterioration of the LC, relate to mild cognitive decline and PD dementia^11–15^. Whether the PD alterations of alpha activity are related to LC degeneration is an open question. Animal models do show that LC firing is related to cortical peri-alpha rhythmic activity^16^. In healthy humans, the amplitude of cortical alpha activity is sensitive to noradrenergic modulation^17^. Further, the pharmacological increase of monoaminergic signalling in healthy adults, including norepinephrine, decreases alpha activity in fronto-central cortices^18^. We therefore hypothesized that LC degeneration in PD is co-expressed in patients with increased, region-specific, alpha activity and pronounced attention impairments.

Rhythmic beta activity (15 – 29 Hz) is also perturbed in PD, and linked to motor deficits^1, 19, 20^, but can be normalized by dopaminergic medications^14, 21, 22^ and by deep brain stimulation of the subthalamic nucleus^22–24^. In the primary motor cortices of early-stage patients with PD, beta activity is increased compared to healthy levels^25^, but this effect reverses as patients reach the moderate stages of the disease^21, 26^. This signifies that in PD, altered dopamine-mediated signalling in the cortico-basal ganglia loop affects frequency-specific, rhythmic cortical activity. Therefore, we asked whether SN degeneration is related to altered expressions of cortical beta-band activity in patients with PD. We hypothesized that loss of neuromelanin in the SN would be related to reduced beta activity and worse motor functions.

Alterations in theta band (5 – 7 Hz) neurophysiological activity have also been shown in PD^27^, though less consistently and rarely in relation to clinical features. More recently, broadband arrhythmic neurophysiological activity has also been shown to shift towards slower frequencies in PD^10, 28–30^, signalling greater relative inhibition^31^ and potentially confounding previously-reported effects reported in low frequency bands^32^. We therefore tested for possible associations between these neurophysiological features, LC and SN neuromelanin, and clinical symptoms.

Ultimately, we tested how associations between neuromelanin depletion, neurophysiological activity, and clinical symptoms map onto the human cortex and relate to the topography of relevant neurochemical systems.

## Methods

### Participants

The Research Ethics Board at the Montreal Neurological Institute reviewed and approved this study. Written informed consent was obtained from every participant following detailed description of the study, and all research protocols complied with the Declaration of Helsinki. Exclusionary criteria for all participants included current neurological (other than PD) or major psychiatric disorder; MEG/MRI contraindications; and unusable neuroimaging or demographic data. Data from patients with mild to moderate (Hoehn and Yahr scale: 1 – 3) idiopathic PD, as diagnosed by a treating neurologist, were included from the Quebec Parkinson Network database (QPN; https://rpq-qpn.ca/)^33^. All patients with PD were prescribed a stable dosage of antiparkinsonian medication with satisfactory clinical response prior to study enrollment. Patients were instructed to take their medication as prescribed before research visits, and thus all data were collected in the practically-defined “on” state.

Magnetoencephalography^34^ data were included for 58 patients with PD who fulfilled the inclusion criteria. Detailed information regarding levodopa medication regimens were available for a subset (N = 23) of these patient participants and used to calculate levodopa equivalent daily dose^35^. Of the 46 patients who also provided information regarding their use of other medications for symptoms of PD, 13 reported taking dopamine agonists. These data were compared to MEG data from a sample of 65 healthy older adults, collated from the Quebec Parkinson Network (N = 10)^33^, PREVENT-AD (N = 40)^36^ and Open MEG Archive (OMEGA; N = 15)^37^ data repositories. These control participants were selected so that their demographic characteristics, including age (Mann-Whitney U test; *W* = 1774.00, *p* = .575), self-reported sex (chi-squared test; χ^2^ = 0.01, *p* = .924), handedness (chi-squared test; χ^2^ = 0.22, *p* = .894), and highest level of education (Mann-Whitney U test; *W* = 1929.00, *p* = .217) did not statistically differ from those of the patient group. All participants underwent the same MEG data collection procedure using the same instrument.

Neuromelanin-sensitive MRI data were available for the same 58 patients with PD who fulfilled the inclusionary criteria and had usable MEG data. Neuromelanin-sensitive MRI data were not available for the healthy controls from the Prevent-AD and OMEGA studies, so neuromelanin-sensitive MRI data from a matched group of 27 healthy older adults from the Quebec Parkinson Network (QPN) was used. These participants underwent the same data collection protocol using the same instrument as the patient group, and were selected so that their demographic characteristics, including age (Mann-Whitney U test; *W* = 892.50, *p* = .303), handedness (chi-squared test; χ^2^ = 0.852, *p* = .653), and highest level of education (Mann-Whitney U test; *W* = 726.50, *p* = .323) did not statistically differ from those of the patient group. The control sample of neuromelanin-sensitive MRI data from the QPN database could not be matched to the patient group on self-reported sex (chi-squared test; χ^2^ = 5.34, *p* = .021), and as such all group-wise neuromelanin-MRI comparisons included sex (in addition to age) as a nuisance covariate.

Group demographic summary statistics and comparisons, as well as clinical summary statistics for the patient group, are provided in Table S1.

### Neuromelanin-sensitive MRI acquisition and processing

Neuromelanin-sensitive MRI data were collected using a 3-Tesla Siemens Prisma scanner with a 32-channel head coil at the Montreal Neurological Institute. Neuromelanin-sensitive sequences were collected using the following parameters: 2D T1 Fast Spin Echo (FSE); echo trains per slice: 60; TR: 600 ms; TE: 10 ms; flip angle: 120°; field of view: 220 mm; slice thickness: 1.8 mm; resolution: 0.7 x 0.7 x 1.8 mm. T1w sequences were collected in the same session using the following parameters: 3D T1 Magnetization Prepared – Rapid Gradient Echo (MPRAGE); TR: 2300 ms; TE: 2.98 ms; flip angle: 9°; field of view: 256 mm; slice thickness: 1 mm; resolution: 1.0 mm isotropic. T1w images underwent denoising, non-uniformity correction, intensity normalization, and registration to stereotaxic space using the NIST Longitudinal Pipeline^38^ and PD126 template^39^ as the registration target.

The neuromelanin-sensitive acquisitions were registered into stereotaxic space using the corresponding linear and non-linear transformations from T1w images processed using the NIST pipeline^38^. Regions of interest including the SN, LC, cerebral peduncles (CP), and pontine tegmentum (PT) were segmented from the processed NM images. SN, CP and PT ROIs were defined manually by an expert on a separate in-house neuromelanin template and were warped to fit the PD126 template. The LC ROI was obtained using a conservative 40% threshold on the Brainstem Navigator probabilistic atlas^40^ aligned on the PD126 template. Segmented ROIs were used to derive the neuromelanin score which represents the integrity of brainstem nuclei (SN, LC) as a ratio, calculated as the normalized average SN NM intensity versus the averaged NM intensity from the CP, and the normalized average LC NM intensity versus the NM intensity from the PT, respectively.

### Clinical & Neuropsychological Testing

Standard clinical assessments were available for most of the patients with PD, including gross motor impairment (Unified Parkinson’s Disease Rating Scale – part III [UPDRS-III]; N = 44)^41^ and general cognitive function (Montreal Cognitive Assessment [MoCA]; N = 51)^42^. In those patients with UPDRS sub-score data available (N = 42), we summed these scores using established criteria^43^ to represent two sets of motor features. *Bradykinesia and rigidity,* but not tremor, are the motor impairments most commonly linked to cortical beta oscillations in PD^20^. We summed the rigidity, finger tapping, pronation/supination of hands, and leg agility sub-scores to represent these features. Axial motor symptoms were also considered in isolation, as they are resistant to administration of levodopa^44^. The following sub-scores of the UPDRS-III were summed to represent *axial* symptoms: speech, facial expression, arising from chair, posture, gait, postural stability, and body bradykinesia. We summed the resting tremor sub-scores to consider tremor symptoms as a potential confound.

Detailed neuropsychological data were available for 49 patients with PD, and as described and validated previously^10, 45^, were used to derive composite scores across five domains: attention (Digit Span – Forward, Backward, and Sequencing; Trail Making Test Part A), executive function (Trail Making Test Part B; Stroop Test – Colors, Words, and Interference; Brixton Spatial Anticipation Test), memory (Hopkins Verbal Learning Test-Revised [HVLT-R] – Learning Trials 1-3, Immediate and Delayed Recall; Rey Complex Figure Test [RCFT] – Immediate and Delayed Recall), language (Semantic Verbal Fluency – Animals & Actions; Phonemic Verbal Fluency – F, A & S; Boston Naming Test), and visuospatial function (Clock Drawing Test – Verbal Command & Copy Command; RCFT – Copy). To utilize as much available data as possible, missing values were excluded pairwise from analysis per each test. Negatively-scored test values were sign-inverted, the data for each individual test were standardized to the mean and standard deviation of the available sample, and these z-scores were then averaged within each domain listed above to derive domain-specific metrics of cognitive function. We focused our analyses on the attention domain *a priori*, due to its established associations with alpha rhythms^46–48^ and noradrenergic function^49–51^. Data regarding disease duration (i.e., years since diagnosis) were also available for N = 46 patients (mean = 6.17 years, SD = 4.94). Additional clinical data are provided in Table S1.

### Magnetoencephalography Data Collection and Analyses

Eyes-open resting-state MEG data were collected using a 275-channel whole-head CTF system (Port Coquitlam, British Columbia, Canada) at a sampling rate of 2400 Hz and with an antialiasing filter with a 600 Hz cut-off. Noise-cancellation was applied using CTF’s software-based built-in third-order spatial gradient noise filters. Recordings lasted a minimum of 5 min^52^ and were conducted with participants in the upright position as they fixated a centrally-presented crosshair. The participants were monitored during data acquisition via real-time audio-video feeds from inside the MEG shielded room, and continuous head position was recorded during all sessions.

MEG preprocessing was performed with *Brainstorm*^53^ unless otherwise specified, with default parameters and following good-practice guidelines^54^. The data were bandpass filtered between 1–200 Hz to reduce slow-wave drift and high-frequency noise, and notch filters were applied at the line-in frequency and harmonics (i.e., 60, 120 & 180 Hz). Signal space projectors (SSPs) were derived around cardiac and eye-blink events detected from ECG and EOG channels using the automated procedure available in *Brainstorm*^55^, reviewed and manually-corrected where necessary, and applied to the data. Additional SSPs were also used to attenuate stereotyped artifacts on an individual basis. Artifact-reduced MEG data were then epoched into non-overlapping 6-s blocks and downsampled to 600 Hz. Data segments still containing major artifacts (e.g., SQUID jumps) were excluded for each session on the basis of the union of two standardized thresholds of ± 3 median absolute deviations from the median: one for signal amplitude and one for its numerical gradient. An average of 78.72 (SD = 14.55; 83.50%) epochs were used for further analysis (patients: 83.14 [SD = 7.65]; controls: 74.77 [SD = 17.82]) and the percent of epochs rejected did not differ between the groups (Mann-Whitney U test; *W* = 1670.00, *p* = .276). Empty-room recordings lasting at least 2 minutes were collected on or near the same day as the participants’ visits and were processed using the same pipeline, with the exception of the artifact SSPs, to model environmental noise statistics for source mapping.

MEG data were coregistered to each individual’s segmented T1-weighted MRI (*Freesurfer recon-all*) using approximately 100 digitized head points. For participants with useable MEG but not MRI data (N_HC_ = 3; N_PD_ = 14), we produced an individualized template with *Brainstorm*, by warping the default *Freesurfer* anatomy to the participant’s head digitization points and anatomical landmarks^56^. We produced source maps of the MEG sensor data with overlapping-spheres head models (15,000 cortical vertices, with current flows of unconstrained orientation) and the dynamic statistical parametric mapping (dSPM) approach, informed by estimates of sensor noise covariance derived from the empty-room MEG recordings.

We obtained vertex-wise estimates of power spectrum density (PSD) from the source-imaged MEG data using Welch’s method (3-s time windows with 50% overlap), which we normalized to the total power of the frequency spectrum at each cortical location. These PSD data were next averaged over all artifact-free 6-s epochs for each participant, and the PSD root-mean-squares across the three orthogonal current flow orientations at each cortical vertex location was projected onto a template cortical surface (*FSAverage*) for comparison across participants.

To separately consider rhythmic versus arrhythmic cortical activity, we parameterized the PSDs with *specparam* (*Brainstorm* Matlab version; frequency range = 2–40 Hz; Gaussian peak model; peak width limits = 0.5 –12 Hz; maximum n peaks = 3; minimum peak height = 3 dB; proximity threshold = 2 standard deviations of the largest peak; fixed aperiodic; no guess weight)^32^ and extracted the vertex-wise exponent of arrhythmic spectral components. The rhythmic (i.e., aperiodic-corrected) spectra were derived by subtracting these arrhythmic components from the original PSDs. Rhythmic components of interest were then extracted by averaging over canonical frequency bands (delta: 2–4 Hz; theta: 5–7 Hz; alpha: 8–12 Hz; beta: 15–29 Hz)^55^. This procedure produced five maps of neurophysiological brain activity per participant: one for rhythmic activity in each of the four canonical frequency bands and one for broadband arrhythmic activity. These maps were regressed on SN and LC neuromelanin scores in whole-cortex analyses (see *Statistical Analyses*, below).

The parameters of the *specparam* rhythmic model fits were also extracted for each vertex location to investigate which accounted for significant relationships. The extracted parameters included the maximum amplitude, frequency-at-maximum-amplitude, and bandwidth (full width at half maximum) of each Gaussian-modeled rhythmic peak. For the alpha-frequency relationships, these features were extracted within the 4 – 15 Hz frequency range (bandwidth extended to account for potential alpha-frequency slowing in PD^57^), and within the 15 – 29 Hz range for beta-frequency relationships. These values were averaged within each participant across the vertices of each significant cluster, excluding any vertices where no rhythmic component was identified. The number of rhythmic peaks detected within the frequency range was summed across all vertices of the relevant cortical cluster, and divided by the total number of possible peaks (i.e., 3 maximum peaks * n cluster vertices) to derive a percentage peak detection probability.

To investigate the possible confounding effects of head motion, eye movements, and heart-rate variability on our MEG data, we extracted the root-sum-square of the reference signals from the MEG head position indicators, EOG, and ECG channels, respectively. None of these metrics significantly differed between patients with PD and controls (Mann-Whitney U tests; head motion: *W* = 1818.00, *p* = .848; EOG: *W* = 1590.00, *p* = .220; ECG: *W* =1898.00, *p* = .715).

### Normative Atlases of Catecholamine Transporter Densities

We used *neuromaps*^58^ to obtain cortical maps of norepinephrine transporter (NET: data from 77 participants using [11C]MRB positron emission tomography) and dopamine transporter (DAT: 174 participants; [123I]-FP-CIT), following previously-established procedures^10, 45, 59^ and parcellated the resulting maps based on both the Desikan-Killiany^60^ and Brainnetome^61^ atlases, to ensure that none of our findings were biased by the choice of atlas.

### Statistical Analyses

Participants with missing data were excluded across analyses pairwise. We used a threshold of *p* < .05 to indicate statistical significance and ran two-tailed tests unless otherwise specified. Significance testing relied on nonparametric models for all cases, to account for any effects of outliers and potential non-normality of the data. All relationships concerning neuromelanin scores in either the LC or SN included the other region as a covariate of no-interest to control for any non-focal effects (e.g., of absolute signal quality or general neurodegeneration).

We derived statistical comparisons across the cortical maps produced, covarying out the effect of age, using *SPM12*. We tested for group differences in neurophysiological features, beyond the effects of age, using independent-samples t-tests. We formulated the relationships between MEG derivatives and neuromelanin scores as multiple regressions per each neurophysiological feature:

*neurophysiological feature ∼ SN_neuromelanin + LC_neuromelanin + nuisance covariate(s)*.

For models where LC neuromelanin was the covariate of interest, age and SN neuromelanin were included as nuisance covariates. Disease duration (i.e., time since diagnosis) was not included in these models, as it did not significantly covary with LC neuromelanin. Disease duration and SN neuromelanin were found to be significantly associated (*r* = -.39, *p*_PERM_ = .007). As such, for models where SN neuromelanin was the covariate of interest, disease duration was also included as a nuisance covariate, alongside age and LC neuromelanin. This limited the participant sample for these models to those patients whose disease duration was known (N = 46). Initial tests used parametric general linear models, with secondary corrections of the resulting *F*-contrasts for multiple comparisons across cortical locations with nonparametric Threshold-Free Cluster Enhancement (TFCE; E = 1.0, H = 2.0; 5,000 permutations)^62^. We applied a final cluster-wise threshold of *p*_FWE_ < .05 to determine statistical significance, and used the TFCE clusters at this threshold to mask the original statistical values (i.e., vertex-wise *F* values) for visualization. We extracted data from the cortical location exhibiting the strongest statistical relationship in each cluster (i.e., the “peak vertex”) for subsequent analysis (e.g., for testing of potential confounds) and visualization.

We derived general linear models to test for univariate group differences and bivariate linear relationships, with nonparametric permutation testing for derivation of frequentist *p*-values using *lmPerm* in *R*. Where appropriate, we assessed Bayesian evidence for null versus alternative models using Bayes Factors (BF) computed with the *BayesFactor* package in *R*. All reported linear relationships survived multiple comparisons corrections across related tests using a Benjamini-Hochberg FDR approach.

We conducted spatial colocalization analyses between group-level statistical maps and neurochemical atlas data from *neuromaps*^58^ using the *cor.test* function in *R* and nonparametric spin-tests with autocorrelation-preserving null models (5,000 Hungarian spins; threshold: *p* < .05)^63^. To facilitate these comparisons, we averaged all relevant neurophysiological and neurochemical data over the 68 parcels of the Desikan-Killiany atlas prior to analysis. We replicated our observation over the 210 regions of the Brainnetome atlas^61^, confirming that our results are not likely to be biased by the cortical parcellation. We obtained group-level statistical maps by regressing region-wise neurophysiological features on covariates of interest, controlling for age. We then extracted for each parcel the resulting unstandardized beta weights for the covariate of interest to represent the spatial topography of unthresholded group-level statistical effects.

### Data Availability

Magnetoencephalography and T1-weighted MRI data used in the preparation of this work are available through the Clinical Biospecimen Imaging and Genetic (C-BIG) repository (https://www.mcgill.ca/neuro/open-science/c-big-repository)^33^, the PREVENT-AD open resource (https://openpreventad.loris.ca/)^36^, and the OMEGA repository (https://www.mcgill.ca/bic/resources/omega)^37^. Normative neurotransmitter density data are available from *neuromaps* (https://github.com/netneurolab/neuromaps)^58^. Neuromelanin-sensitive MRI data will be made openly available in the future, and until then is available upon reasonable request.

## Results

### Atypical Cortical Neurophysiology and Brainstem Neuromelanin Depletion in Parkinson’s Disease

We found expressions of atypical neurophysiological activity in patients with PD compared to healthy older adults in bilateral posterior (theta band; PD > HC; cluster *p*_FWE_ < .001; peak vertex = x: 33, y: -45, z: -11; Figure 1a, top), precentral (alpha band; PD > HC; cluster *p*_FWE_ = .011; peak vertex = x: -43, y: 14, z: 41; Figure 1a, middle), and occipito-temporal cortices (slope of the aperiodic spectrum; PD < HC; cluster *p*_FWE_ = .013; peak vertex = x: 48, y: -64, z: 25; Figure 1a, bottom). These findings align with previous literature^27, 30^. As expected in patients “on” dopaminergic medications that normalize beta-band activity in PD^14, 21, 22^, we did not find significant group differences in beta rhythmic activity. As such, any observed relationships with beta rhythmic activity should be interpreted with caution, as they do not necessarily reflect a pathological process.

**Figure 1.**
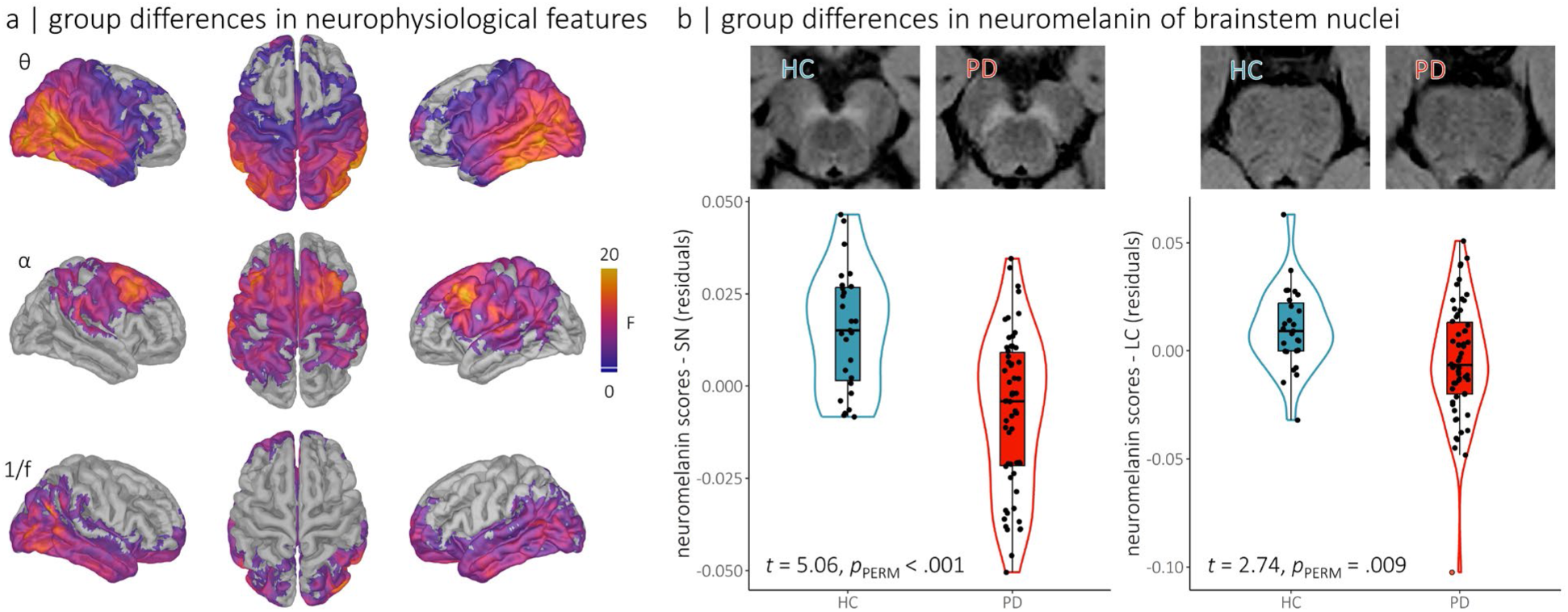
Alterations of regional cortical neurophysiology and neuromelanin depletion of brainstem nuclei in Parkinson’s disease. (a) Cortical maps represent regional clusters of differences in rhythmic (i.e., theta and alpha oscillations) and arrhythmic (i.e., aperiodic 1/f slope) neurophysiological features between patients with Parkinson’s disease and healthy older adults. Colors indicate the strength of the statistical effect (in *F* values), thresholded based on the cluster limits identified using threshold-free cluster enhancement (*p*_FWE_ < .05). No significant differences were observed in the delta and beta bands. (b) Top: representative neuromelanin-sensitive MRI data focused on the substantia nigra (left panel) and the locus coeruleus (right panel) for one participant with Parkinson’s disease (PD, in red) and one age-matched healthy control participant (HC, in blue). Bottom: group differences in neuromelanin scores for substantia nigra (left panel) and locus coeruleus (right panel), with individual points representing participants. Boxes-and-whiskers indicate the median, upper and lower quartiles, and minima/maxima for each group, and violin plots show the associated density distributions.

We also found reduced neuromelanin MRI signals in both the SN (*t* = 5.06, *p*_PERM_ < .001) and the LC (*t* = 2.74, *p*_PERM_ = .009; Figure 1b) of patients. It is important to note that the overlap between MEG and neuromelanin MRI data was predominantly within the PD patient group. Consequently, we employed the data from healthy older adults solely to identify the neuroimaging features that differentiate PD patients from non-affected individuals. All analyses that connected these two types of data (MEG and neuromelanin MRI) were exclusively conducted within the PD patient cohort.

**Figure 2.**
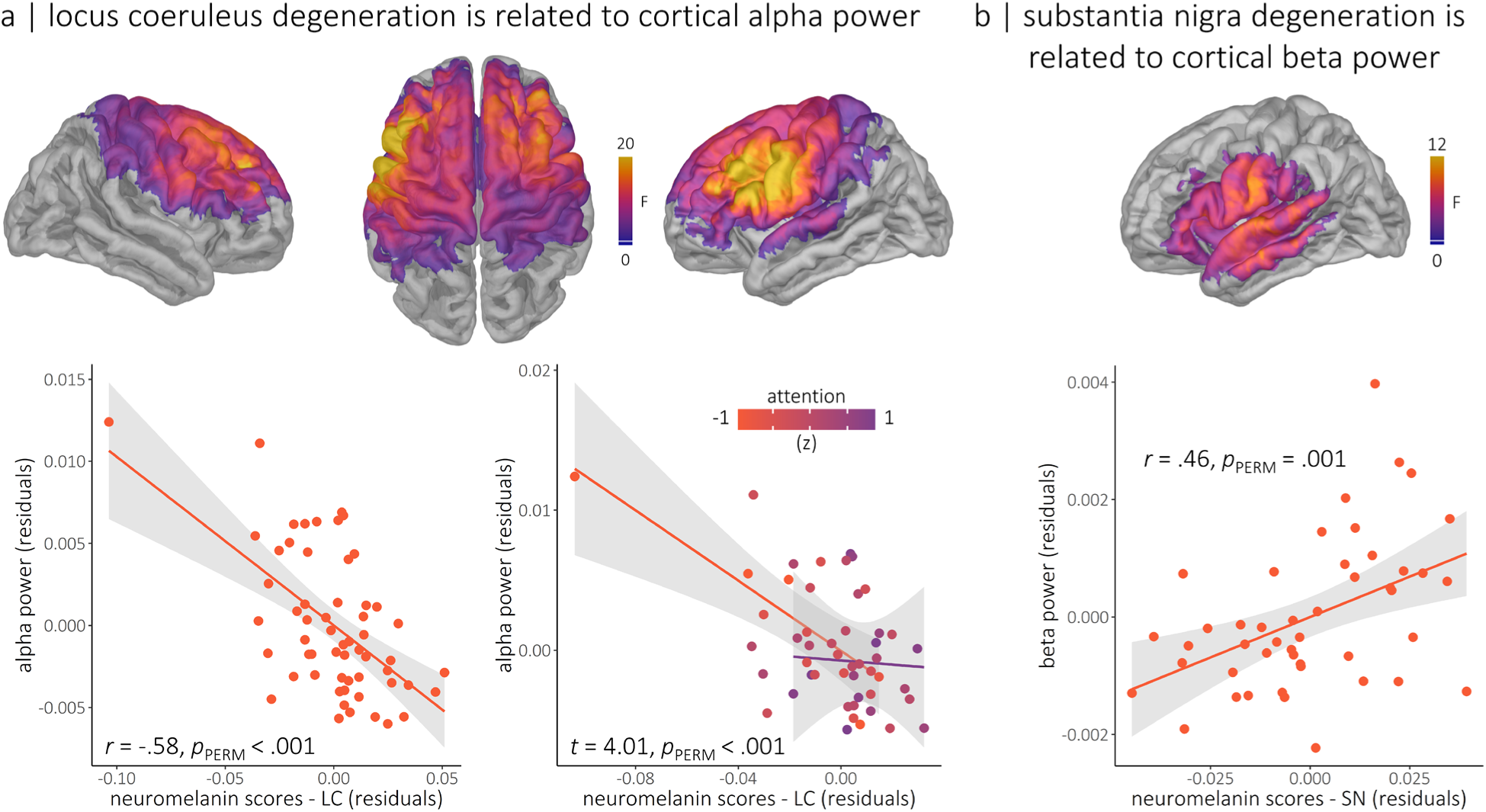
Regional cortical rhythmic neurophysiological features related to brainstem nuclei neuromelanin. (a) The maps show the cortical regions where locus coeruleus neuromelanin scales with rhythmic alpha power, beyond the effects of substantia nigra neuromelanin and age. The nature of this relationship is displayed in scatter plots of peak-vertex alpha power values on the bottom left, with the partial correlation coefficient and permuted p-value overlaid. The line plots on the bottom right reflect the significant moderation of this relationship by attention abilities (indicated by colors of lines; orange = impaired attention, purple = preserved attention), such that individuals with worse attentional impairments exhibited a stronger relationship between locus coeruleus degeneration and rhythmic alpha power. (b) The map shows the cortical regions where substantia nigra neuromelanin scores are associated with rhythmic beta power, beyond the effects of locus coeruleus neuromelanin, disease duration, and age. The nature of this relationship is displayed in the scatter plot below using peak-vertex beta power values, with the partial correlation coefficient and permuted p-value overlaid. Colors on the cortical maps indicate the strength of the statistical effect (in F values), thresholded based on the cluster limits identified using threshold-free cluster enhancement (*p_FWE_* < .05). We used nonparametric permutation testing to account for the influence of outliers in these associations. Also note that the subjective outlier in the top left of the scatterplots in panel (a) did not exert undue influence on the model (defined as a Cook’s distance > 3 SD from the group mean).

### Depletion of Neuromelanin in Locus Coeruleus and Substantia Nigra is Associated with Atypical Cortical Neurophysiology in Parkinson’s Disease

We found that depletion of neuromelanin in the LC is associated with increased alpha activity in the bilateral fronto-motor cortices of patients (cluster *p_FWE_* = .006; peak *p_PERM_* < .001; peak vertex = x: -52, y: 7, z: 34; Figure 2a), beyond the effects of neuromelanin depletion in the SN. This relationship is contributed by the presence (i.e., peak likelihood; r = -.38, *p_PERM_* < .001; post-hoc tests) and amplitude (r = -.30, *p_PERM_* = .018) of the alpha peak in the frequency spectrum of regional activity (Figure S1), not by other spectral parameters (center frequency: *p_PERM_* = .583; BF_01_ = 3.19; width: *p_PERM_* = .961; BF_01_ = 3.36; all-features model versus frequency-features model: BF_10_ = 5.40). Note that this relationship remains significant (*t* = -3.86, *p_PERM_* < .001) even after the exclusion of a subjective outlier data point (see orange data point in Figure 1b). This point did not exert undue influence on the model (Cook’s distance = .047; 1.47 SD from the group mean), and thus it could not be excluded based on any empirical threshold. Furthermore, no significant hemispheric lateralization of this effect was observed (laterality index; *t* = -0.25, *p*_PERM_ = .706). This association between LC neuromelanin and alpha activity is more pronounced in patients with stronger attention impairments (moderating effect: *t* = 4.01, *p*_PERM_ < .001; Figure 2a, bottom right). Here too, the subjective outlier point did not exert undue influence on the model (Cook’s distance = .014; -0.18 SD from the group mean).

We found that the association of alpha activity with LC neuromelanin is stronger in the bilateral fronto-motor cortex, where alpha activity also scales negatively with attention scores (r = .69, pSPIN < .001; Figure 3a) and which features the highest concentrations of norepinephrine transporters (NET; r = -.65, *p_SPIN_* < .001; Figure 3b). Both of these alignments were replicated using a different parcellation (i.e., Brainnetome; attention: r = .64, *p_SPIN_* < .001; NET: r = -.65, *p_SPIN_* < .001).

**Figure 3.**
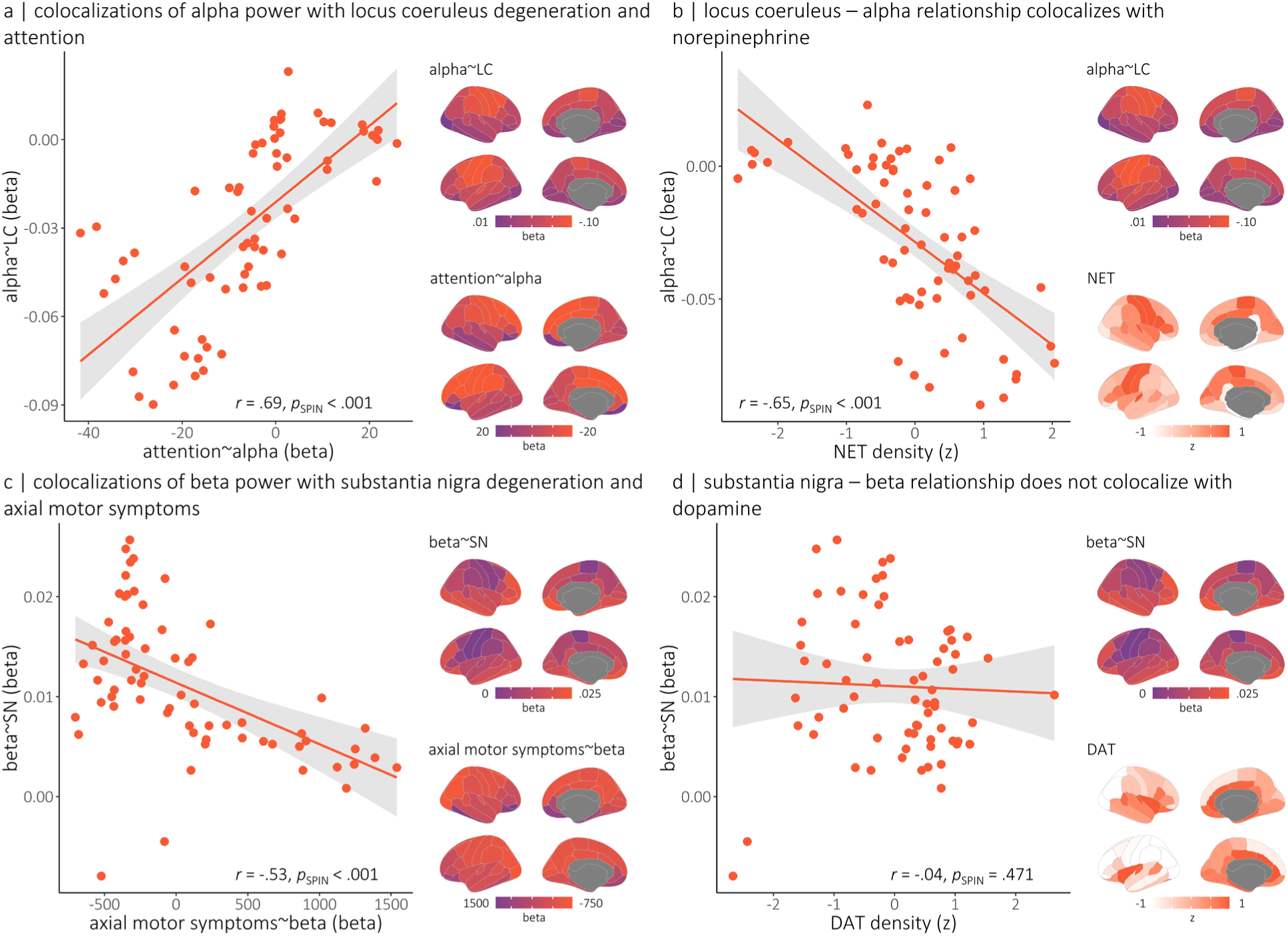
Colocalization of cortical associations between rhythmic neurophysiological features, clinical symptoms and brainstem nuclei neuromelanin. (a) Cortical maps indicate the region-wise linear relationships, in unstandardized regression (i.e., beta) weights, between alpha power and locus coeruleus neuromelanin (top) and attention (bottom). The scatter plot to the left indicates the alignment between these maps (each dot is from a parcel of the Desikan-Killiany cortical atlas), with r--values and p-values overlaid. (b) Colocalization of the region-wise alpha-LC relationships shown in (a) with the density of cortical norepinephrine transporter. (c) Similar to the effects shown in (a), but concerning relationships between rhythmic beta power, substantia nigra neuromelanin, and axial motor symptoms. (d) Similar to the effects shown in (b), but concerning colocalization of the region-wise beta-SN relationships with the density of cortical dopamine transporter. Note that null distributions for estimating p-values were generated using 5,000 autocorrelation-preserving spatial permutations of the data.

Our data also show that depletion of neuromelanin in the SN is associated with decreased beta activity in the left lateral cortices of patients (cluster *p*_FWE_ = .010; peak *p*_PERM_ = .001; peak vertex = x: -64, y: -7, z: 28; Figure 2b), with the strongest peak in the postcentral gyrus. This relationship is significant beyond the effects of disease duration and neuromelanin depletion in the LC. This association is specific to the amplitude of beta activity (*r* = .41, *p*_PERM_ = .006; Figure S1), and is not related to other peak parameters (peak likelihood, *p*_PERM_ = .150; BF_01_ = 0.96; center frequency: *p*_PERM_ = .804; BF_01_ = 2.77; width: *p*_PERM_ = .725; BF_01_ = 2.97; all-features model versus frequency-features and detection probability model: BF_10_ = 6.61). This effect is not significantly moderated by motor symptoms (i.e., summed sub-scores from the UPDRS-III; *axial*: p_PERM_ = .667, *bradykinesia and rigidity*: p_PERM_ = .128).

The cortical topography of the association between beta activity and SN neuromelanin is aligned with that of the association of regional beta activity with axial (*r* = .69, *p*_SPIN_ < .001; Figure 3c) but not bradykinesia and rigidity (*r* = -.01, *p*_SPIN_ = .543) motor symptoms. This alignment was also replicated using a different parcellation (i.e., *Brainnetome*; *r* = .52, *p*_SPIN_ < .001). There was no significant alignment of the beta-SN relationship with the cortical atlas of dopamine transporters (DAT; *r* = -.04, *p*_SPIN_ = .471; Figure 3d).

These effects were not substantively affected by additional nuisance covariates (i.e., disease duration, eye movements, heart rate variability, head motion, and number of accepted trials; all *p*’s remained < .01). Neither the alpha-LC nor the beta-SN relationship was significantly moderated by the use of dopamine agonists (alpha-LC: *p*_PERM_ = .482; beta-SN: *p*_PERM_ = .623) or the levodopa equivalent daily dose (alpha-LC: *p*_PERM_ = .667; beta-SN: *p*_PERM_ = .076), and both of these relationships remained significant when the severity of resting-tremor symptoms was included as a nuisance covariate (alpha-LC: N = 42, *t* = -3.77, *p*_PERM_ < .001; beta-SN: N = 40, *t* = 2.61, *p*_PERM_ = .007).

## Discussion

Neurophysiological changes in patients with PD affect several aspects of the electrophysiological spectrum and relate to the cognitive and motor features of the disease^10, 12, 27, 28, 45, 64^. Reductions in beta-band activity are the most well-studied in PD, with associations to the severity^1, 19, 20^ and eventual amelioration^14, 21–24^ of motor impairments. Increased alpha-band cortical rhythms have received less attention, but are thought to relate to cognitive impairments and progression to PD dementia^11–15^.

Herein, we report, using a new multimodal approach, that alpha- and beta-band cortical neurophysiology are differentially associated with PD degeneration of neuromelanin-rich cells in brainstem nuclei. Our data show that beta activity relates to loss of dopaminergic neurons in the SN, and alpha activity to the depletion of noradrenergic cells in the LC. The cortical regions where alpha activity is the most strongly related to LC neuromelanin also have higher concentrations of norepinephrine transporter. We found that the cortical regions of strong association between alpha activity and LC neuromelanin are also those where alpha activity scales with the severity of attentional impairments. We also found that in regions where regional beta activity is related to SN neuromelanin, stronger beta activity also signals more severe axial motor impairments.

These findings suggest dissociable norepinephrine-alpha-cognitive and dopamine-beta-motor pathophysiological pathways in PD. The relationship between LC degeneration and alpha activity is particularly significant from a pathophysiological standpoint, as it establishes a new link between previous lines of research showing similar associations between noradrenergic dysfunction^3^, atypical expressions of alpha activity^11–15^ and cognition in PD, as well as between norepinephrine and alpha activity in the healthy adult brain^16–18^. We also report associations between beta activity, SN degeneration, and motor impairments, thereby confirming previous, albeit scattered, observations^1^. Our results provide converging evidence for the association of dopaminergic nigrostriatal dysfunction in motor-related beta changes in PD. Furthermore, these findings broaden the existing literature by highlighting levodopa-resistant associations between motor impairments and beta rhythmic activity in lateral peri-central cortices. It should be noted, however, that these beta-SN relationships were weaker than their alpha-LC counterparts, and we did not find significant differences in amplitude of cortical beta activity between patients with PD and control participants. Further, the beta-SN effects we observed did not map to the primary motor cortices. This may be attributable to the patients being under their normal regimen of dopaminergic medications when they participated in the study. These medications ameliorate motor symptoms^65^ and normalize beta activity^14, 21^, which is likely to reduce the meaningful variability to be modeled in our study, and therefore the size of the association effects. This possibility is supported by the trending moderation of the beta-SN relationship by levodopa equivalent daily dose: those patients taking a higher dose of dopamine therapy showed a qualitative reduction in the beta-SN effect. Additionally, only the axial motor symptoms that are resistant to levodopa treatment^44^ were found to be associated with the same beta oscillations that scaled with SN neuromelanin depletion. The relationship between SN neuromelanin and beta activity was not stronger in cortical regions of high DAT density. However, dopaminergic projections from the SN synapse more prominently with the striatum than directly with the neocortex^66^. Given that reduced dopaminergic signaling from the SN affects cortical activity indirectly, it is less probable that cortical dopamine systems significantly modulate the relationship between beta activity and SN degeneration. Nonetheless, the absence of data from patients in their “off” medication state constitutes a significant limitation of our study, especially concerning the relationships observed with beta rhythmic activity. In contrast, out findings suggest relatively strong evidence that the association between LC neuromelanin and alpha rhythmic activity is mediated by noradrenergic mechanisms, which are unlikely to be influenced by levodopa administration.

We also emphasize that few studies so far^10, 29–31, 67, 68^ have considered the respective associations of rhythmic and arrhythmic neurophysiological activity with PD. Yet, pathological changes in rhythmic versus arrhythmic electrophysiology call for distinct mechanistic interpretations. For instance, the arrhythmic slope of the neurophysiological power spectrum is steeper in patients with PD, indicating a broadband reduction in excitatory-versus-inhibitory firing^31, 69^. This arrhythmic shift was present in our participants but did not significantly relate to degeneration of the SN nor LC. Instead, we found that only rhythmic spectral activity in the alpha and beta bands was related to the integrity of brainstem nuclei. This reinforces previous conceptualizations of PD as affecting primarily rhythmic neurophysiological activity^64^. We also found that these relationships are replicated only with the presence (i.e., detection probability) and strength (i.e., peak amplitude) of band-limited rhythmic activity in fronto-motor cortices, rather than its frequency definitions (i.e., peak frequency or bandwidth).

Theta band activity also differed between patients with PD and healthy older adults, but did not significantly relate to neuromelanin levels in the LC nor SN. It is possible that these neurophysiological alterations are instead associated with other types of neurochemical depletions in PD, such as acetylcholine^70^. Alternatively, theta band effects may be secondary to the primary neuropathological hallmarks of PD (e.g., Lewy bodies). Future studies investigating in more detail theta band activity in PD are therefore necessary.

In sum, the present study advances the notion of a noradrenergic basis of atypical expressions of regional alpha rhythmic activity in patients with PD, with relevance for attention functions; it also strengthens current theoretical constructs of the role of dopaminergic dysfunction in the changes of rhythmic beta activity related to motor functions in those patients. Future research will substantiate the causal nature of these relationships with longitudinal study designs and, e.g., non-invasive neuromodulation approaches. These findings are potentially translatable as a biomarker of treatment response in PD: neurophysiology would be expected to respond more dynamically to disease-modifying therapeutics than measures of brainstem nucleus integrity, and thus may be valuable within the relatively short time-frame of clinical trials. The present results also hint at the possibility that clinical interventions combining noradrenergic pharmacotherapies with frequency-targeted neurostimulation may be advantageous in PD. Overall, this study advances our understanding of the neurochemical bases of neurophysiological alterations in patients with PD, of which non-dopaminergic mechanisms remain particularly underexplored.

## Data Availability

Magnetoencephalography and T1-weighted MRI data used in the preparation of this work are available through the Clinical Biospecimen Imaging and Genetic (C-BIG) repository (https://www.mcgill.ca/neuro/open-science/c-big-repository), the PREVENT-AD open resource (https://openpreventad.loris.ca/), and the OMEGA repository (https://www.mcgill.ca/bic/resources/omega). Normative neurotransmitter density data are available from neuromaps (https://github.com/netneurolab/neuromaps). Neuromelanin-sensitive MRI data will be made openly available in the future, and until then is available upon reasonable request.

## Acknowledgments

This work was supported to AIW by a Banting Postdoctoral Fellowship from the Canadian Institutes of Health Research (CIHR; BPF-186555) and grant F32-NS119375 from the United States National Institutes of Health (NIH); to EAF as a Foundation Grant from the Canadian Institutes of Health Research (CIHR; FDN-154301) and the CIHR Canada Research Chair (Tier 1) of Parkinson’s Disease; to DLC from a Rapid Response Grant from the Weston Family Foundation (RR171117); and to SB from a NSERC Discovery grant (RGPIN-2020-06889), a grant from the Healthy Brains for Healthy Lives initiative of McGill University under the Canada First Research Excellence Fund and the CIHR Canada Research Chair (Tier 1; CRC-2017-00311) for Neural Dynamics of Brain Systems, and a grant from the NIH (R01-EB026299). Data collection and sharing for this project was provided by the Quebec Parkinson Network (QPN), the Pre-symptomatic Evaluation of Novel or Experimental Treatments for Alzheimer’s Disease (PREVENT-AD; release 6.0) program, and the Open MEG Archives (OMEGA). The funders had no role in study design, data collection and analysis, decision to publish, or preparation of the manuscript.

The QPN is funded by a grant from Fonds de recherche du Québec - Santé (FRQS) and the Canada First Research Excellence Fund, awarded through the HBHL initiative at McGill University. PREVENT-AD was launched in 2011 as a $13.5 million, 7-year public-private partnership using funds provided by McGill University, the FRQS, an unrestricted research grant from Pfizer Canada, the Levesque Foundation, the Douglas Hospital Research Centre and Foundation, the Government of Canada, and the Canada Fund for Innovation. Private sector contributions are facilitated by the Development Office of the McGill University Faculty of Medicine and by the Douglas Hospital Research Centre Foundation (http://www.douglas.qc.ca/). OMEGA and the Brainstorm app are supported by funding to SB from the NIH (R01-EB026299), a Discovery grant from the Natural Science and Engineering Research Council of Canada (436355-13), and the CIHR Canada research Chair in Neural Dynamics of Brain Systems (CRC-2017-00311).

## Author Contributions

Alex I Wiesman: conceptualization, methodology, formal analysis, writing – original draft, visualization, funding acquisition; Victoria Madge: methodology, formal analysis, writing – review & editing; Edward A Fon: conceptualization, resources, writing – review & editing, supervision, project administration, funding acquisition; Alain Dagher: conceptualization, resources, writing – review & editing, supervision, project administration, funding acquisition; D Louis Collins: conceptualization, resources, writing – review & editing, supervision, project administration, funding acquisition; Sylvain Baillet: conceptualization, resources, writing – review & editing, supervision, project administration, funding acquisition.

## Declaration of Interests

The authors declare no competing interests, financial or otherwise.

## Notes

### Competing Interest Statement

The authors have declared no competing interest.

### Author Declarations

The Research Ethics Board at the Montreal Neurological Institute reviewed and approved this study.

### Summary of Updates

Updated to fix formatting errors.

## References

1. Jenkinson N, Brown P. New insights into the relationship between dopamine, beta oscillations and motor function. Trends in neurosciences 2011;34:611–618.

2. Braak H, Del Tredici K, Rüb U, De Vos RA, Steur ENJ, Braak E. Staging of brain pathology related to sporadic Parkinson’s disease. Neurobiology of aging 2003;24:197–211.

3. Del Tredici K, Braak H. Dysfunction of the locus coeruleus–norepinephrine system and related circuitry in Parkinson’s disease-related dementia. Journal of Neurology, Neurosurgery & Psychiatry 2013;84:774–783.

4. Aston-Jones G, Rajkowski J, Cohen J. Locus coeruleus and regulation of behavioral flexibility and attention. Progress in brain research 2000;126:165–182.

5. Smith A, Nutt D. Noradrenaline and attention lapses. Nature 1996.

6. Sasaki M, Shibata E, Tohyama K, et al. Neuromelanin magnetic resonance imaging of locus ceruleus and substantia nigra in Parkinson’s disease. Neuroreport 2006;17:1215–1218.

7. Sulzer D, Cassidy C, Horga G, et al. Neuromelanin detection by magnetic resonance imaging (MRI) and its promise as a biomarker for Parkinson’s disease. NPJ Parkinson’s disease 2018;4:11.

8. McCusker MC, Wiesman AI, Spooner RK, et al. Altered neural oscillations during complex sequential movements in patients with Parkinson’s disease. NeuroImage: Clinical 2021;32:102892.

9. da Silva Castanheira J, Wiesman A, Hansen J, et al. Neurophysiological brain-fingerprints of motor and cognitive decline in Parkinson’s disease. medRxiv 2023:2023.2002. 2003.23285441.

10. Wiesman AI, da Silva Castanheira J, Degroot C, et al. Adverse and compensatory neurophysiological slowing in Parkinson’s disease. Progress in Neurobiology 2023:102538.

11. Wiesman AI, Donhauser PW, Degroot C, et al. Aberrant neurophysiological signaling underlies speech impairments in Parkinson’s disease. medRxiv 2022.

12. Wiesman AI, Heinrichs-Graham E, McDermott TJ, Santamaria PM, Gendelman HE, Wilson TW. Quiet connections: Reduced fronto-temporal connectivity in nondemented Parkinson’s Disease during working memory encoding. Hum Brain Mapp 2016.

13. Schmiedt C, Meistrowitz A, Schwendemann G, Herrmann M, Basar-Eroglu C. Theta and alpha oscillations reflect differences in memory strategy and visual discrimination performance in patients with Parkinson’s disease. Neuroscience letters 2005;388:138–143.

14. Bosboom J, Stoffers D, Stam C, et al. Resting state oscillatory brain dynamics in Parkinson’s disease: an MEG study. Clinical Neurophysiology 2006;117:2521–2531.

15. Dubbelink KTO, Stoffers D, Deijen JB, Twisk JW, Stam CJ, Berendse HW. Cognitive decline in Parkinson’s disease is associated with slowing of resting-state brain activity: a longitudinal study. Neurobiology of aging 2013;34:408–418.

16. Berridge CW, Foote SL. Effects of locus coeruleus activation on electroencephalographic activity in neocortex and hippocampus. Journal of Neuroscience 1991;11:3135–3145.

17. Dahl MJ, Mather M, Werkle-Bergner M. Noradrenergic modulation of rhythmic neural activity shapes selective attention. Trends in cognitive sciences 2022;26:38–52.

18. Albrecht MA, Roberts G, Price G, Lee J, Iyyalol R, Martin-Iverson MT. The effects of dexamphetamine on the resting-state electroencephalogram and functional connectivity. Human Brain Mapping 2016;37:570–588.

19. Heinrichs-Graham E, Wilson TW, Santamaria PM, et al. Neuromagnetic evidence of abnormal movement-related beta desynchronization in Parkinson’s disease. Cerebral cortex 2014;24:2669–2678.

20. Little S, Brown P. The functional role of beta oscillations in Parkinson’s disease. Parkinsonism & related disorders 2014;20:S44–S48.

21. Heinrichs-Graham E, Kurz MJ, Becker KM, Santamaria PM, Gendelman HE, Wilson TW. Hypersynchrony despite pathologically reduced beta oscillations in patients with Parkinson’s disease: a pharmaco-magnetoencephalography study. Journal of Neurophysiology 2014;112:1739–1747.

22. Giannicola G, Marceglia S, Rossi L, et al. The effects of levodopa and ongoing deep brain stimulation on subthalamic beta oscillations in Parkinson’s disease. Experimental neurology 2010;226:120–127.

23. Abbasi O, Hirschmann J, Storzer L, et al. Unilateral deep brain stimulation suppresses alpha and beta oscillations in sensorimotor cortices. Neuroimage 2018;174:201–207.

24. Quinn EJ, Blumenfeld Z, Velisar A, et al. Beta oscillations in freely moving Parkinson’s subjects are attenuated during deep brain stimulation. Movement Disorders 2015;30:1750–1758.

25. Pollok B, Krause V, Martsch W, Wach C, Schnitzler A, Südmeyer M. Motor-cortical oscillations in early stages of Parkinson’s disease. The Journal of physiology 2012;590:3203–3212.

26. Vardy AN, van Wegen EE, Kwakkel G, Berendse HW, Beek PJ, Daffertshofer A. Slowing of M1 activity in Parkinson’s disease during rest and movement–an MEG study. Clinical Neurophysiology 2011;122:789–795.

27. Boon LI, Geraedts VJ, Hillebrand A, et al. A systematic review of MEG-based studies in Parkinson’s disease: The motor system and beyond. Human brain mapping 2019;40:2827–2848.

28. Wiesman AI, Donhauser PW, Degroot C, et al. Aberrant neurophysiological signaling associated with speech impairments in Parkinson’s disease. npj Parkinson’s Disease 2023;9:61.

29. Wang Z, Mo Y, Sun Y, et al. Separating the aperiodic and periodic components of neural activity in Parkinson’s disease. European Journal of Neuroscience 2022;56:4889–4900.

30. Helson P, Lundqvist D, Svenningsson P, Vinding MC, Kumar A. Cortex-wide topography of 1/f-exponent in Parkinson’s disease. bioRxiv 2023:2023.2001. 2019.524792.

31. Wiest C, Torrecillos F, Pogosyan A, et al. The aperiodic exponent of subthalamic field potentials reflects excitation/inhibition balance in Parkinsonism. Elife 2023;12:e82467.

32. Donoghue T, Haller M, Peterson EJ, et al. Parameterizing neural power spectra into periodic and aperiodic components. Nature neuroscience 2020;23:1655–1665.

33. Gan-Or Z, Rao T, Leveille E, et al. The Quebec Parkinson network: a researcher-patient matching platform and multimodal biorepository. Journal of Parkinson’s disease 2020;10:301–313.

34. Baillet S. Magnetoencephalography for brain electrophysiology and imaging. Nature neuroscience 2017;20:327.

35. Schade S, Mollenhauer B, Trenkwalder C. Levodopa equivalent dose conversion factors: an updated proposal including opicapone and safinamide. Movement disorders clinical practice 2020;7:343.

36. Tremblay-Mercier J, Madjar C, Das S, et al. Open Science Datasets from PREVENT-AD, a Longitudinal Cohort of Pre-symptomatic Alzheimer’s Disease. NeuroImage: Clinical 2021:102733.

37. Niso G, Rogers C, Moreau JT, et al. OMEGA: the open MEG archive. Neuroimage 2016;124:1182–1187.

38. Aubert-Broche B, Fonov VS, García-Lorenzo D, et al. A new method for structural volume analysis of longitudinal brain MRI data and its application in studying the growth trajectories of anatomical brain structures in childhood. Neuroimage 2013;82:393–402.

39. Madge V, Fonov VS, Xiao Y, et al. A dataset of multi-contrast unbiased average MRI templates of a Parkinson’s disease population. Data in Brief 2023:109141.

40. Bianciardi M, Toschi N, Edlow BL, et al. Toward an in vivo neuroimaging template of human brainstem nuclei of the ascending arousal, autonomic, and motor systems. Brain connectivity 2015;5:597–607.

41. Goetz CG, Tilley BC, Shaftman SR, et al. Movement Disorder Society-sponsored revision of the Unified Parkinson’s Disease Rating Scale (MDS-UPDRS): scale presentation and clinimetric testing results. Movement disorders: official journal of the Movement Disorder Society 2008;23:2129–2170.

42. Nasreddine ZS, Phillips NA, Bédirian V, et al. The Montreal Cognitive Assessment, MoCA: a brief screening tool for mild cognitive impairment. J Am Geriatr Soc 2005;53:695–699.

43. Stebbins GT, Goetz CG. Factor structure of the Unified Parkinson’s Disease Rating Scale: motor examination section. Movement disorders: official journal of the Movement Disorder Society 1998;13:633–636.

44. Bejjani B-P, Gervais D, Arnulf I, et al. Axial parkinsonian symptoms can be improved: the role of levodopa and bilateral subthalamic stimulation. Journal of Neurology, Neurosurgery & Psychiatry 2000;68:595–600.

45. Wiesman AI, da Silva Castanheira J, Fon EA, Baillet S, Group P-AR, Network QP. Structural and neurophysiological alterations in Parkinson’s disease are aligned with cortical neurochemical systems. medRxiv 2023:2023.2004. 2004.23288137.

46. Klimesch W. α-band oscillations, attention, and controlled access to stored information. Trends Cogn Sci 2012;16:606–617.

47. Wiesman AI, Groff BR, Wilson TW. Frontoparietal Networks Mediate the Behavioral Impact of Alpha Inhibition in Visual Cortex. Cereb Cortex 2018.

48. Wiesman AI, Wilson TW. Alpha Frequency Entrainment Reduces the Effect of Visual Distractors. Journal of cognitive neuroscience 2019:1–12.

49. Unsworth N, Robison MK. A locus coeruleus-norepinephrine account of individual differences in working memory capacity and attention control. Psychonomic bulletin & review 2017;24:1282–1311.

50. Gabay S, Pertzov Y, Henik A. Orienting of attention, pupil size, and the norepinephrine system. Attention, Perception, & Psychophysics 2011;73:123–129.

51. Beane M, Marrocco R. Norepinephrine and acetylcholine mediation of the components of reflexive attention: implications for attention deficit disorders. Progress in neurobiology 2004;74:167–181.

52. Wiesman AI, da Silva Castanheira J, Baillet S. Stability of spectral estimates in resting-state magnetoencephalography: Recommendations for minimal data duration with neuroanatomical specificity. Neuroimage 2022;247:118823.

53. Tadel F, Baillet S, Mosher JC, Pantazis D, Leahy RM. Brainstorm: a user-friendly application for MEG/EEG analysis. Computational intelligence and neuroscience 2011;2011.

54. Gross J, Baillet S, Barnes GR, et al. Good practice for conducting and reporting MEG research. Neuroimage 2013;65:349–363.

55. Niso G, Tadel F, Bock E, Cousineau M, Santos A, Baillet S. Brainstorm pipeline analysis of resting-state data from the open MEG archive. Frontiers in neuroscience 2019;13:284.

56. Tadel F, Bock E, Niso G, et al. MEG/EEG group analysis with brainstorm. Frontiers in neuroscience 2019;13:76.

57. Stoffers D, Bosboom J, Deijen J, Wolters EC, Berendse H, Stam C. Slowing of oscillatory brain activity is a stable characteristic of Parkinson’s disease without dementia. Brain 2007;130:1847–1860.

58. Markello RD, Hansen JY, Liu Z-Q, et al. Neuromaps: structural and functional interpretation of brain maps. BioRxiv 2022.

59. Hansen JY, Shafiei G, Markello RD, et al. Mapping neurotransmitter systems to the structural and functional organization of the human neocortex. Nature Neuroscience 2022.

60. Desikan RS, Ségonne F, Fischl B, et al. An automated labeling system for subdividing the human cerebral cortex on MRI scans into gyral based regions of interest. Neuroimage 2006;31:968–980.

61. Fan L, Li H, Zhuo J, et al. The human brainnetome atlas: a new brain atlas based on connectional architecture. Cerebral cortex 2016;26:3508–3526.

62. Smith SM, Nichols TE. Threshold-free cluster enhancement: addressing problems of smoothing, threshold dependence and localisation in cluster inference. Neuroimage 2009;44:83–98.

63. Markello RD, Misic B. Comparing spatial null models for brain maps. NeuroImage 2021;236:118052.

64. Oswal A, Brown P, Litvak V. Synchronized neural oscillations and the pathophysiology of Parkinson’s disease. Current opinion in neurology 2013;26:662–670.

65. Poewe W, Seppi K, Tanner CM, et al. Parkinson disease. Nature reviews Disease primers 2017;3:1–21.

66. Haber SN. The place of dopamine in the cortico-basal ganglia circuit. Neuroscience 2014;282:248–257.

67. Darmani G, Drummond NM, Ramezanpour H, et al. Long-Term Recording of Subthalamic Aperiodic Activities and Beta Bursts in Parkinson’s Disease. Movement Disorders 2023;38:232–243.

68. Kroesche M, Kannenberg S, Butz M, et al. Slowing of Frontocentral Beta Oscillations in Atypical Parkinsonism. bioRxiv 2022.

69. Gao R, Peterson EJ, Voytek B. Inferring synaptic excitation/inhibition balance from field potentials. Neuroimage 2017;158:70–78.

70. Calabresi P, Picconi B, Parnetti L, Di Filippo M. A convergent model for cognitive dysfunctions in Parkinson’s disease: the critical dopamine–acetylcholine synaptic balance. The Lancet Neurology 2006;5:974–983.

