## Supplementary Materials for "Associations between neuromelanin depletion and cortical rhythmic activity in Parkinson’s disease"

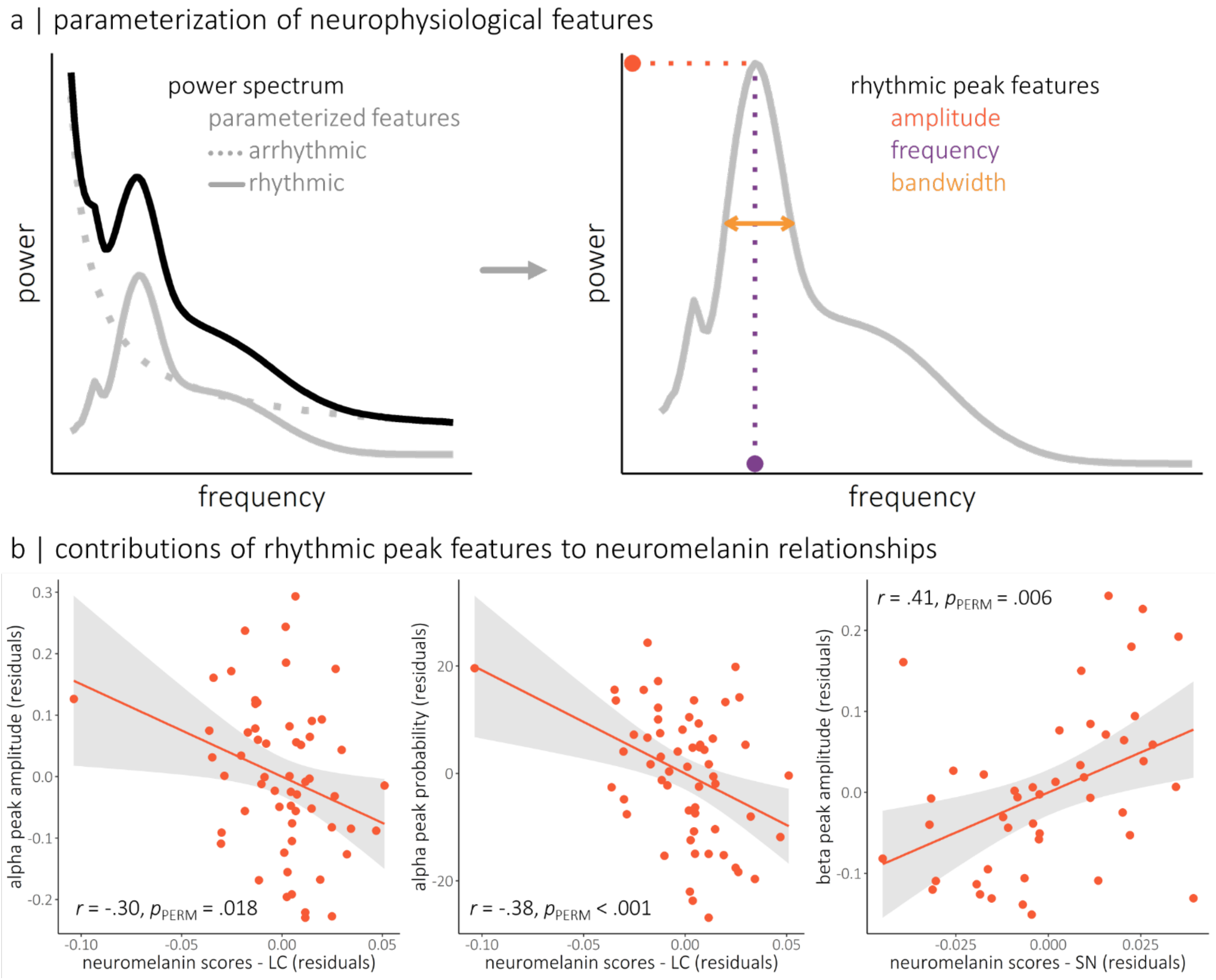


**Figure S1. Parameterization of neurophysiological features and their contributions to the observed neuromelanin-neurophysiology relationships.** (a) The conceptual figure to the left indicates the parameterization of power spectra (black line) into arrhythmic (dotted grey line) and rhythmic (solid grey line) features. From the rhythmic model, several features of each peak can be derived (right), including the peak amplitude (orange), peak frequency (purple), and peak bandwidth (i.e., at full width at half maximum; gold). The probability of detecting a rhythmic peak can also be quantified, to represent the relative presence/absence of detectable oscillatory activity around a given frequency. (b) The alpha and beta rhythmic peak features were collapsed spectrally (i.e., alpha: 4 – 15 Hz; beta: 15 – 29 Hz) and spatially (i.e., over the clusters shown in Figure 2), and modeled against LC (for alpha) and SN (for beta) neuromelanin scores, to determine which rhythmic features were contributing significantly to the reported neuromelanin-neurophysiology relationships. Associated *r*-values and permuted *p*-values for each comparison are overlaid. Shaded intervals indicate 95% confidence intervals. Note that nonparametric permutation testing was used to account for any influence of oultiers on these estimated linear relationships.

**Table S1.** Demographic comparisons and patient group clinical profile.

| Group | Age (years) | Sex (% male) | Handedness (# left/ambi) | Education (years) |
|---|---|---|---|---|
| **$HC_{MEG}$** (N = 65) | 63.02 (8.13)$^{\#}$ | 64.62$^{\#}$ | 3/3$^{\#}$ | 15.85 (3.72)$^{‡\#}$ |
| **$HC_{NM}$** (N = 27) | 65.82 (9.83)$^{\#}$ | 37.04* | 1/0$^{‡‡‡\#}$ | 14.91 (4.78)$^{‡‡\#}$ |
| **PD** (N = 58) | 64.22 (8.37) | 63.79 | 2/2 | 14.66 (2.45) |

| Group | MoCA (N = 51) | UPDRS-III (N = 44) | Hoehn & Yahr (N = 44) | % Taking DA Agonists (N = 46) |
|---|---|---|---|---|
| **PD** | | | | |
| **Range** | 12 – 30 | 7 – 71 | 1 – 3 | – |
| **Mean** (SD) | 24.43 (4.08) | 32.80 (15.20) | 1.92 (0.74) | 28.30 |

$HC_{MEG}$: healthy control group – magnetoencephalography data; $HC_{NM}$: healthy control group – neuromelanin-sensitive magnetic resonance imaging data; PD: Parkinson's disease group; MoCA: Montreal Cognitive Assessment; UPDRS-III: Unified Parkinson's Disease Rating Scale part III; DA: Dopamine. Unless otherwise indicated, values indicate means and associated parentheticals indicate standard deviations. $^{‡}$N = 52; highest level of education was not reported by six $HC_{MEG}$ participants. $^{‡‡}$N = 22; highest level of education was not reported by 5 $HC_{NM}$ participants. $^{‡‡‡}$N = 14; handedness was not reported by 13 $HC_{NM}$ participants. $^{\#}$not significant, *$p < .05$; derived from between-groups tests relative to the PD group using Mann-Whitney U tests and chi-square tests for continuous and categorical variables, respectively.
